# Downregulation of hippocampal activity improves memory performance in individuals at risk of Alzheimer’s disease

**DOI:** 10.1101/2025.10.14.25333311

**Authors:** Katharina Klink, Yves Lysser, Marina Wunderlin, Piotr Radojewski, Charlotte Teunissen, Michael Orth, Jessica Peter

**Affiliations:** University Hospital of Old Age Psychiatry and Psychotherapy, University of Bern, Bern, Switzerland; Translational Imaging Centre, Swiss Institute for Translational and Entrepreneurial Medicine, Bern, Switzerland; Graduate School for Health Sciences, University of Bern, Bern, Switzerland; University Institute of Diagnostic and Interventional Neuroradiology, Inselspital, Bern University Hospital, University of Bern, Switzerland; Neurochemistry Laboratory, Department of Clinical Chemistry, Vrije Universiteit Amsterdam, Amsterdam, Netherlands

**Keywords:** AD biomarkers, RCT, p-Tau181, blood-based biomarker

## Abstract

**INTRODUCTION:** In people at risk of Alzheimer’s disease (AD), hippocampal hyperactivity relates to worse hippocampus-dependent task performance and promotes AD pathology. Reducing hippocampal hyperactivity promises to improve hippocampus-dependent memory performance.

**METHODS:** 78 participants (69.29±5.87 years old, 60% female) used real-time fMRI neurofeedback twice to downregulate the hippocampus or a control region. We assessed hippocampal activity and memory performance before and after neurofeedback. We classified AD pathology risk using a blood-based biomarker, an established risk-score, or clinical characterisation.

**RESULTS:** During hippocampus, but not during control-region neurofeedback, individuals significantly downregulated hippocampal activity with the effect still detectable one week later. In those at high risk of AD pathology, with all three classifications, stronger hippocampal downregulation resulted in stronger memory improvement, independent of baseline hippocampal hyperactivity.

**DISCUSSION:** Downregulating hippocampal activity using fMRI neurofeedback may be a complementary treatment for people at risk for AD, well before there is hippocampal hyperactivity.

## Background

In Alzheimer’s disease (AD), the hippocampus, among other regions, degenerates with an accumulation of extra-cellular beta amyloid (Aβ) plaques and intra-cellular phosphorylated tau tangles. The degenerative process begins a long time before the emergence of clinical signs and before a diagnosis of mild cognitive impairment (MCI) can be made [1]. Biomarkers, for instance from cerebrospinal fluid (CSF), positron emission tomography (PET), or blood may help identify people at risk of developing AD pathology before hippocampal dependent cognition becomes affected [1]. Pattern separation, for instance, is an ability that relies heavily on the hippocampus [2]. When the hippocampus degenerates, such as in patients with MCI, performance in pattern separation tasks declines. Despite worse task performance, however, several fMRI studies reported *higher* activity in the hippocampus in patients with MCI compared to healthy individuals [2–4]. The biological processes that lead to hyperactivity in the hippocampus are not fully understood. Data from rodent AD models indicate that inhibitory interneurons within the hippocampus decline. This shifts the excitatory–inhibitory balance towards over-excitation, thus unbalancing the promotion and suppression of neuronal firing. This could be relevant for the neuropathological processes underlying AD pathogenesis as neuronal excitability increased near amyloid plaques and in hippocampal neurons containing neurofibrillary tangles. Neuronal hyperactivity then promoted amyloid accumulation and tau release in a vicious cycle [5]. In humans, recurrent collaterals within a subregion of the hippocampus seem to become disinhibited, leading to unconstrained activation of auto-associative connections (i.e., neurons that connect to other neurons within this region, including back to themselves) [6]. The accumulation of amyloid beta may then inhibit glutamate reuptake with subsequent loss of inhibitory receptors as well as synapses – and hence to hyperactivity [6]. In later disease stages, the accumulation of tau tangles promotes the loss of dendritic spines in neurons, with subsequent hypoactivation and progressive neurodegeneration [7]. An initial hyperactivity therefore ultimately leads to neuronal loss and to hypoactivation, as has been observed in patients with AD [8].

The phase with hippocampal hyperactivity in the disease trajectory may present an opportunity for interventions aiming to reduce it. If successful, this could improve hippocampal function and slow down the biological processes that advance because of hippocampal hyperactivity (Supplementary Fig. 1) [2]. It has repeatedly been found, for example, that low-dose antiepileptic drugs reduced hippocampal hyperactivity and improved pattern separation performance [2,3,9]. Pharmacological interventions, however, entail a risk of side effects. In addition, an elderly population is likely to be on other medication, so - apart from possible side effects - drug interactions may be a problem. An alternative approach may be real-time fMRI neurofeedback, during which individuals learn to regulate region-specific brain activity. This is accomplished by measuring and processing brain activity changes in targeted brain regions in real time and by providing feedback to an individual [10]. Neurofeedback has been employed successfully to reduce hyperactivity in other regions, such as the amygdala or posterior cingulate cortex patients with post-traumatic stress disorder, leading to symptom improvement [11,12]. In the current study, we therefore investigated whether real-time fMRI neurofeedback can reduce hippocampal hyperactivity and improve pattern separation performance in individuals at risk of AD.

## Methods

### Participants and clinical characterisation

Recruitment and data acquisition including follow-up assessments took place from July 2020 until December 2023. We screened *n* = 223 participants for study inclusion that were recruited via advertisement or flyers circulated in Bern. Of these, *n* = 77 did not sign up and *n* = 59 did not meet inclusion criteria. The remaining *n* = 87 participants were included in the study^1^. Of those, *n* = 9 were lost to follow-up. Hence, *n* = 78 were finally analysed (30 male, 48 female; 69.29 ± 5.87 years old). Since there is evidence to suggest that amyloid pathology and hippocampal hyperactivity are related [14], we used three different approaches to assess amyloid positivity risk in our participants (Fig. 1). First, we calculated that risk by using plasma phosphorylated tau 181 (p-Tau181), a blood-based biomarker that differentiated amyloid positive from amyloid negative individuals in clinical samples (confirmed either by CSF or PET) and that was most predictive of future progression to AD [15]. Second, we used a formula to calculate the risk of amyloid positivity in all participants based on a model provided by Palmqvist and colleagues [16]. This model is based on age, sex, verbal episodic memory performance, and *APOE* genotype. Finally, we based calculations on clinical diagnosis (i.e., cognitively healthy vs. amnestic MCI) using established criteria [17,18]. Participants were classified as having amnestic MCI if they fulfilled criteria for a diagnosis of ‘amnestic MCI due to AD pathology’ according to established criteria [17]. That required an impairment in the delayed recall of a previously learned list of words (1.5 standard deviations below age-, gender, and education-adjusted norms). In addition, they needed to a) report a cognitive complaint, b) show no impairment in activities of daily living, c) no dementia but d) signs of neuronal injury (i.e., hippocampal volume loss or medial temporal lobe atrophy) [18]. Participants were considered cognitively healthy if their Montreal Cognitive Assessment (MoCA) score was ≥ 26 [19]. All data were collected at the University of Bern. All participants provided written informed consent before the study. The cantonal Ethics Committee approved the study (2019-00958) which was done in accordance with the Declaration of Helsinki. The study was registered with clinicaltrials.gov (NCT04020744).

**Figure 1.**
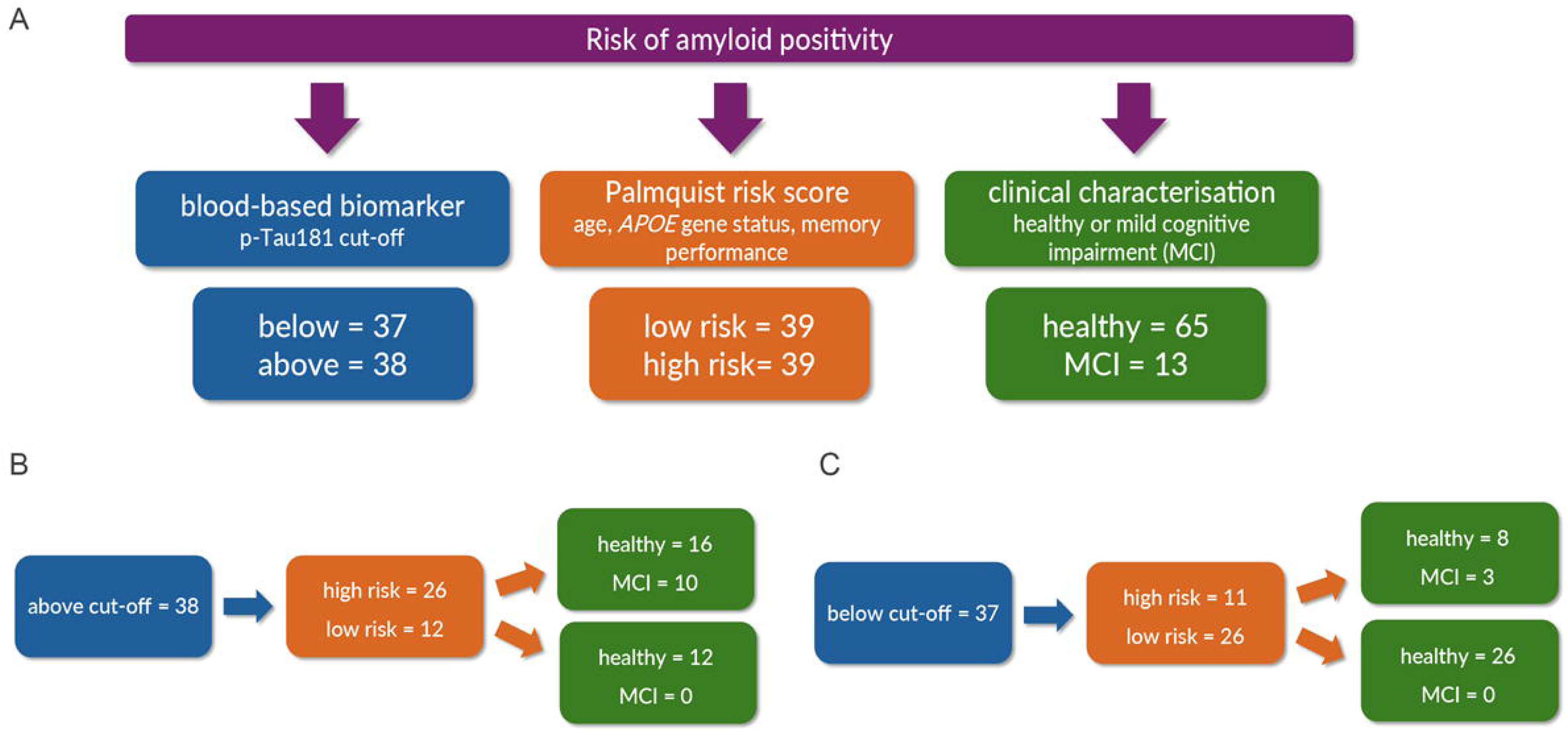
Classification of amyloid positivity risk in our participants. **(A)** We classified the risk of amyloid positivity using three independent methods. The overlap between those methods is shown in **(B)** for participants above the p-Tau181 cut-off and in **(C)** for participants below that cut-off.

### Study Procedure

We assigned participants to one of two groups in a single-blind, randomized, and parallel-group design (simple randomization; ratio 1:3). One group downregulated activity in the hippocampus. The other group downregulated activity in a control region (i.e., regions surrounding the intraparietal sulcus which are not involved in pattern separation). A person not involved in data collection (JP) generated the allocation sequence using a Matlab Script and assigned participants to the intervention groups. All participants attended five times, with 1-2 weeks between each assessment (Fig. 2A). At the first assessment, we drew blood and evaluated cognitive performance. At the second assessment, we acquired structural and functional MRI data, including an fMRI pattern separation task, and we assessed verbal episodic memory performance [20]. At the third and fourth assessment, all participants downregulated brain activity using real-time fMRI neurofeedback. The fifth assessment was identical to the second without acquiring structural MRI data.

**Figure 2.**
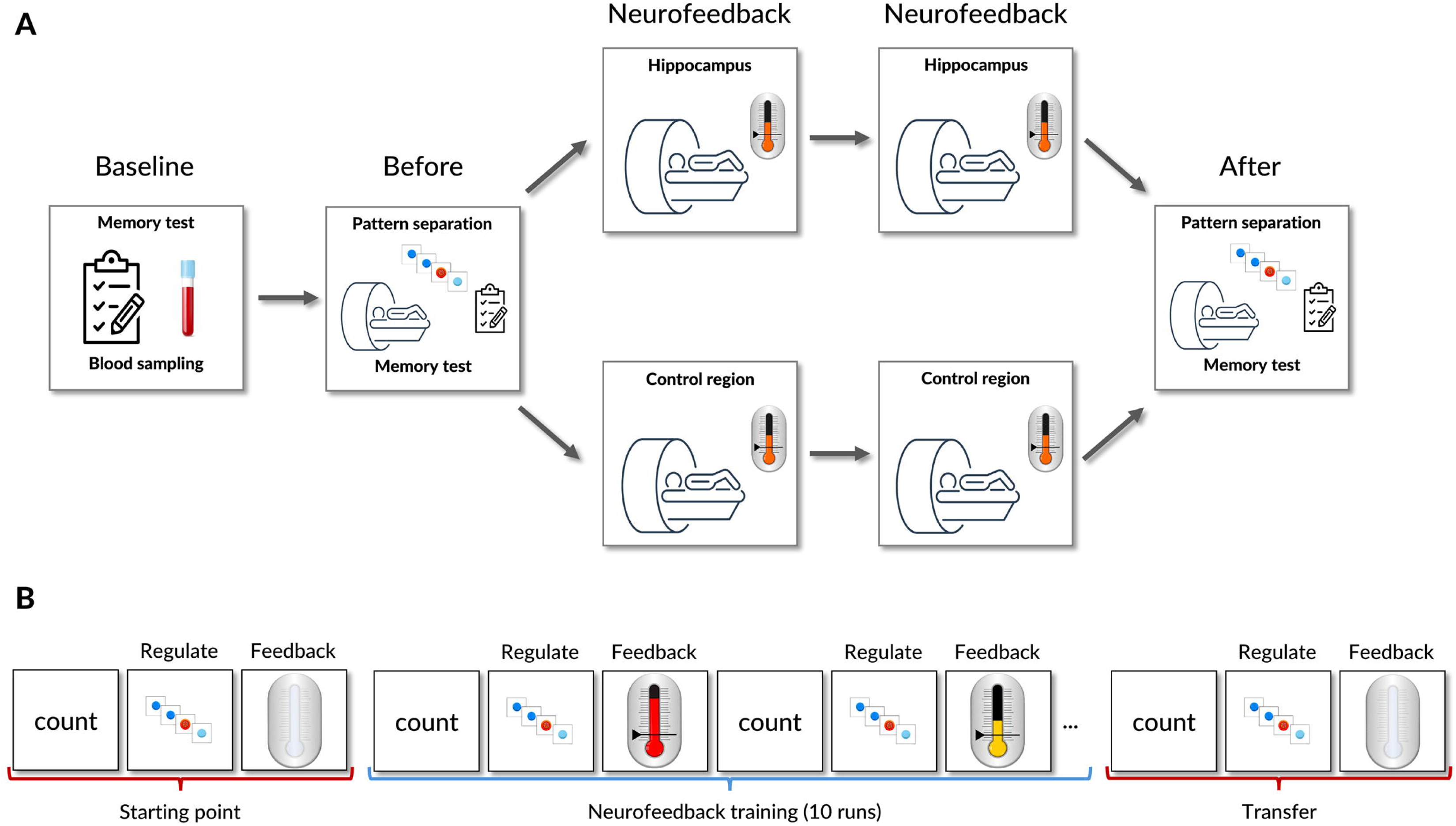
Study procedures with neurofeedback. **A)** At baseline, we tested verbal episodic memory and collected blood from all participants. Before neurofeedback, we examined a pattern separation task using fMRI as well as verbal episodic memory again. Then, two sessions of neurofeedback followed in which participants downregulated either hippocampal activity or activity in a control region. After neurofeedback, we assessed pattern separation performance again as well as verbal episodic memory. All examinations were around one week apart. **B)** Each neurofeedback session included a starting point and a transfer run, in which no feedback was provided to the participants as well as 10 runs in which feedback was provided.

### Neurofeedback intervention

Detailed information regarding the neurofeedback intervention can be found in our study protocol [21]. In brief, two neurofeedback sessions followed the initial baseline assessment, with ten neurofeedback runs in total (Fig. 2A and B). Each session lasted 1 hour and contained five runs during which participants received feedback about activity changes in their hippocampus or the control region. We asked participants to reduce brain activity by using mental strategies of their choice. Participants were not aware of the brain region they were about to downregulate (i.e., single-blind study). Each session began with a starting point assessment and ended with a transfer run, during which participants were asked to regulate brain activity but received no feedback (Fig. 2B). At the starting point, participants were, most likely, not aware which strategies regulated their brain activity. At the end of each session, after receiving feedback for an hour, they presumably learned which strategies regulated their brain activity. Hence, we compared the starting point to the transfer run to examine whether participants learned how to downregulate activity. We used a blocked design that consisted of counting backwards, regulating brain activity, and receiving feedback (Fig. 2B). During regulation, we used an implicit pattern separation task that elicits hippocampal activity but does not require any response from participants [22]. Feedback was provided using a thermometer icon ranging from red (high activity) to green (low activity). Before moving into the scanner, we asked participants to explain the task in their own words to ensure correct understanding. After each session, we debriefed participants to find out which strategies they used for downregulation.

### Explicit pattern separation task

Around 1 week before and around 1 week after neurofeedback, we assessed pattern separation performance in all participants using an explicit version of a pattern separation task (explicit = responses are provided by button presses). We used a three-alternative forced choice task, in which participants viewed new, old, or similar colour images of common objects. We asked them to indicate by button press whether objects were new (i.e., novel items), old (i.e., repeated items), or similar (i.e., lure items). Of critical interest were participants’ responses on lure (i.e., similar) items. A response of ‘old’ to those items indicates a bias towards pattern completion, while responding ‘similar’ to a similar item indicates efficient pattern separation. All participants completed two subsequent runs of the task, with each run including 120 new objects, 40 old objects, and 40 similar objects. 32 old and 32 similar items were shown with a lag of 2 to 12 items (mean = 5.9 items) while another 8 old and 8 similar items were presented with a 0-lag. In total, we presented 400 images for 2 s with an inter-stimulus interval of 0.5 s. Similar to previous studies, we used the lure discrimination index (LDI) for statistical analysis. The LDI tracks the ability to remember the rich details of an item in order to correctly identify it as ‘similar’ and not endorse it as ‘old’ [23]. In addition, we used the number of correctly identified similar items as well as the number of errors (i.e., similar items incorrectly classified as ‘old’) for statistical analysis.

### Magnetic resonance imaging

We acquired MRI data on a 3T Magnetom Prisma scanner (Siemens Healthineers, Erlangen, Germany) using a 32-channel head coil. For structural MRI, we used the following parameters in an MP2RAGE sequence: 176 slices, repetition time (TR) = 5000 ms, echo time (TE) = 2.98 ms, inversion time (TI) = 2500 ms, flip angle = 5°, matrix = 240 × 256, voxel size = 1x 1 x 1 mm. For functional MRI, we acquired data using a T2*-weighted EPI sequence (1000 volumes for pattern separation; 160 volumes for neurofeedback) with the following parameters: flip angle = 30 °, TR = 1000 ms for pattern separation/TR = 2000 ms for neurofeedback, TE = 37 ms, matrix size = 92 × 92, voxel size = 2.5 × 2.5 × 2.5 mm. We presented the task using PsychoPy (version 3.1, [24]) and Celeritas Fiber Optic Response System (Psychology Software Tools, PA, USA).

We processed T1-weighted images using the default options of CAT12 (version 2170, https://neuro-jena.github.io/cat/) running on MATLAB 2019a (MathWorks, Sherborn, MA). In brief, this included skull-stripping, tissue classification, and spatial normalization using DARTEL registration [25]. Quality checks included visual screening of normalized bias corrected volumes and checking data homogeneity when correcting for total intracranial volume and age. We evaluated hippocampal volume using the percentage of total intracranial volume, corrected for head size [26]. In addition, an experienced neuroradiologist (PR) rated medial temporal lobe atrophy (MTA) [27]. We pre-processed functional MRI data using SPM12 (version 7771, https://www.fil.ion.ucl.ac.uk/spm/software/spm12/) running on MATLAB 2019a. This included realignment, co-registration to the structural image, normalisation, and smoothing with a 6 mm full-width at half maximum (FWHM) Gaussian kernel. To remove low frequency noise from pre-processed data, a high-pass filter was applied to the data using SPM12’s default settings. Before smoothing, we included normalized functional images (MNI), normalized white matter, and cerebrospinal fluid segments in a principal component analysis (PCA) using PhysIO toolbox [28]. The PCA resulted in six components, which we used as noise regressors for first-level analysis in addition to six motion parameters.

### Blood-based biomarkers

We collected non-fasted EDTA blood through venipuncture. Samples were centrifuged for 10 min at 1800 rpm at room temperature. Plasma was then aliquoted in ≤ 0.5 mL-portions in polypropylene tubes and stored at −80°C until dry-ice transportation to the Neurochemistry Laboratory at Amsterdam University. There, before analysis, samples were shortly thawed at room temperature and centrifuged at 10’000 g for 10 min. Subsequently, phosphorylated Tau181 (p-Tau181) levels were measured on a Simona HDX system (Qanterix, Billerica, USA) according to the manufacturer’s instructions and with four-times in-board sample dilution. We focused on p-Tau181 as a recent study revealed that it was more sensitive than amyloid beta 42/40 ratio for detecting amyloid positivity (as confirmed using PET or CSF) in healthy people and in patients with MCI [15]. In addition, p-Tau181 was sensitive for the detection of future progression to AD [29]. We additionally derived *APOE* genotypes from blood.

### Statistical analysis

We first characterized participants before neurofeedback took place. We examined whether hippocampal volume, medial temporal lobe atrophy [30], hippocampal activity, and behavioural task performance differed between individuals above or below the p-Tau181 cut- off of amyloid positivity, between individuals with high or low amyloid risk based on a model provided by Palmquist and colleagues, or between healthy participants and patients with amnestic MCI. For behavioural task performance before neurofeedback, we used the fMRI pattern separation task and calculated the lure discrimination index (LDI) [23]. This index tracks the ability to remember the rich details of items so that one can differentiate similar items from new items. It is usually the ability in which patients with amnestic MCI perform worse [31]. In addition, we used the free recall of a verbal episodic memory task to assess behavioural task performance.

We then went on to examine whether participants significantly downregulated activity in the hippocampus during neurofeedback. We calculated the difference in activity between the starting point and the transfer run (i.e., beginning and end of neurofeedback in which no feedback was provided, Fig. 2). Then, we examined whether the regulation of neuronal activity in the hippocampus or the control region influenced hippocampal activity or behavioural performance in the fMRI pattern separation task one week after neurofeedback. We used the LDI as well as the percentage of items that were correctly identified as ‘similar’ and the percentage of errors (i.e., similar items that were classified as ‘old’) as has been done in previous studies [2,3,9,32]. Finally, we examined whether a change in hippocampal activity was correlated with performance change in the pattern separation task.

For statistical analysis, we classified participants in three ways: based on a p-Tau181 cut-off for amyloid positivity (i.e., above or below), based on a model provided by Palmquist and colleagues (i.e., low or high amyloid risk), or based on clinical characterisation (i.e., cognitively healthy vs. amnestic MCI).

### Behavioural analysis

We used t-tests or Χ^2^-tests with group (above or below p-Tau181 cut-off, high or low amyloid risk, healthy or patients with amnestic MCI) as between-subject variable and hippocampal volume, medial temporal lobe atrophy, hippocampal activity, LDI, or episodic memory performance as dependent variables for the characterization of our participants before neurofeedback took place.

To examine behavioural performance change after neurofeedback, we used linear mixed models. We included a by-subject random intercept for repeated measurements (before or after neurofeedback) and the LDI, correct identification of similar items as well as errors as dependent variables. To examine whether functional activity change was associated with behavioural performance change, we used Pearson correlations. We corrected for multiple comparisons using Bonferroni’s method.

### fMRI analysis

For first level analysis of neurofeedback, we fitted general linear models for regulation performance during the starting point or transfer run, i.e., before and after ten neurofeedback training runs in two consecutive neurofeedback sessions. Each model included regressors for ‘no regulation’, ‘regulation’, and ‘feedback’ blocks. In addition, we included six motion and noise regressors in each model. The noise regressors were used for an image-based correction to account for physiological artifacts, similar to previous neurofeedback studies [33]. Our contrast of interest was ‘regulation’ – ‘no regulation’. For second-level analysis, we extracted individual beta estimates based on that contrast for the hippocampus using two-sample *t*-tests. For first level analysis of the explicit pattern separation task, we fitted general linear models for pattern separation performance one week before and one week after neurofeedback. Each model included regressors for correct or incorrect responses to new objects, old objects, and similar objects. We included six motion and noise regressors in each model. Our contrast of interest was ‘correct similar’ – ‘correct new’ responses, as in previous studies [2,4]. For second-level analysis, we extracted individual beta estimates based on that contrast for the hippocampus using two-sample *t*-tests.

We then entered the individual mean contrast estimates into a linear mixed effect model [34,35]. We included a by-subject random intercept for the repeated measurement (before or after neurofeedback) and added age as well as hippocampal volume as covariates.

## Results

### Classification of study participants

We classified participants in three ways. First, we used a plasma p-Tau181 cut-off derived from previous studies in which it was used to predict amyloid positivity (confirmed by amyloid PET or in CSF [15]). Of the 78 participants, 38 were above the blood-based biomarker cut-off (Fig. 1; Supplementary Table 1, Supplementary Fig. 2). Second, we used a formula to calculate the risk of amyloid positivity from age, sex, *APOE* gene status, and verbal episodic memory performance [16]. Of the 78 participants, 39 had a high risk of amyloid positivity (Fig. 1; Supplementary Table 2, Supplementary Fig. 3). Third, participants were classified as having amnestic MCI using established criteria (19). Of the 78 participants, 13 fulfilled criteria for amnestic MCI (Fig. 1; Supplementary Table 3, Supplementary Fig. 4). Classification of participants as having a high risk of developing Alzheimer’s disease using any of these methods was associated with less hippocampal grey matter (p-Tau cut-off: *p* = .001; amyloid positivity risk: *p* = .003; MCI: *p* < .001), more medial temporal lobe atrophy (p-Tau cut-off: *p* = .009; amyloid positivity risk: *p* = .020; MCI: *p* < .001), and worse verbal episodic memory performance (p-Tau cut-off: *p* = .004; amyloid positivity risk: *p* < .001; MCI: *p* < .001). In addition, they performed worse in the pattern separation task (p-Tau cut- off: *p* = .044; amyloid positivity risk: *p* = .006; MCI: *p* = .001) despite higher hippocampal activity (p-Tau cut-off: *p* = .086; amyloid positivity risk: *p* = .227; MCI: *p* = .033) before neurofeedback (Fig. 3).

**Figure 3.**
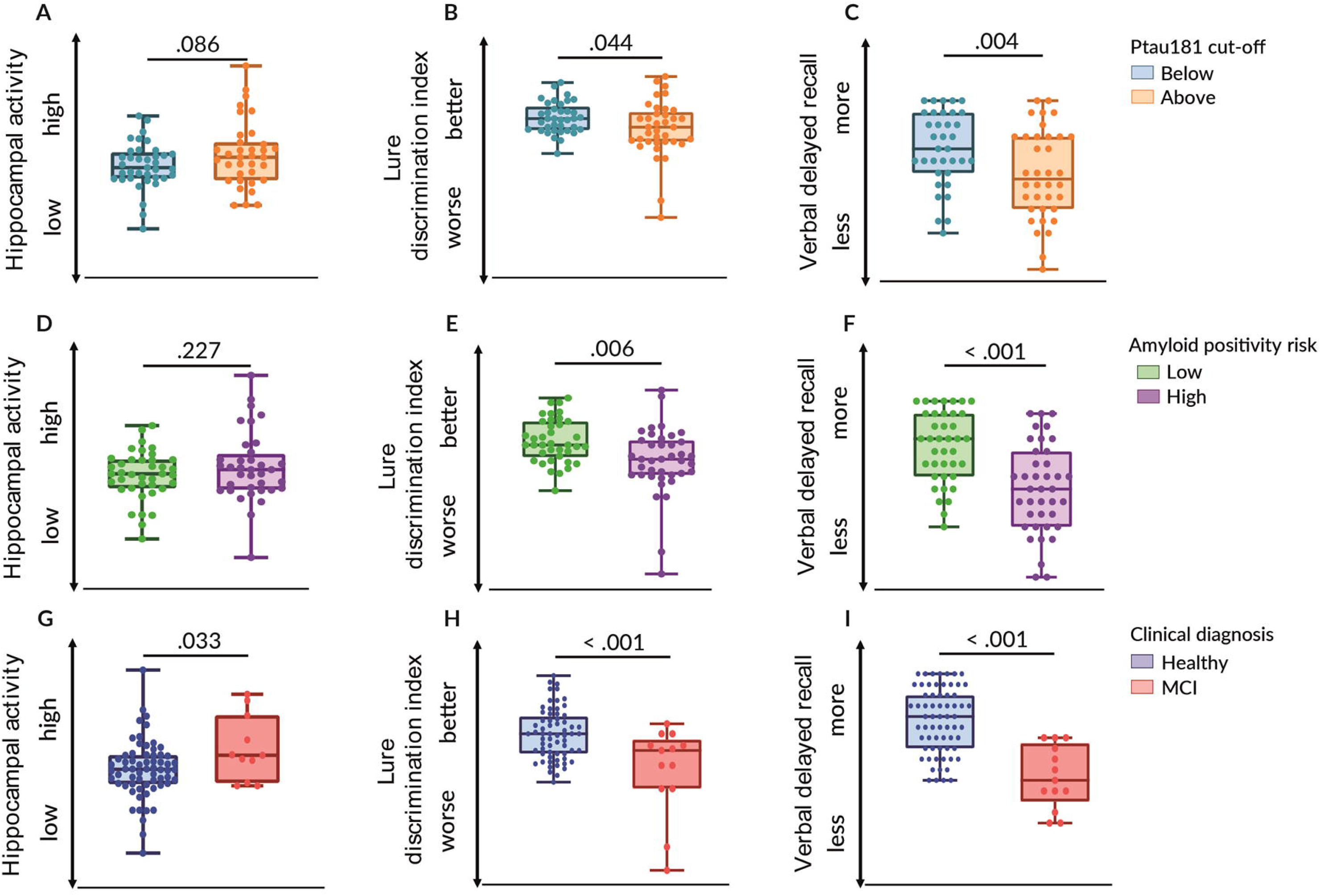
Hippocampal activity and behavioural task performance before neurofeedback. We split the sample based on **(A-C)** a blood-based biomarker (i.e., p- Tau181), **(D-F)** amyloid positivity risk calculation, or **(G-H)** clinical diagnosis. Error bars indicate standard error of the mean. Boxplots indicate min to max values.

### Neurofeedback from the hippocampus reduced hippocampal activity

Two neurofeedback sessions followed the initial baseline assessment, with ten neurofeedback runs in total (Fig. 2A and B). Participants either downregulated activity in the hippocampus or in a control region (i.e., regions surrounding the intraparietal sulcus). We calculated the difference in activity between a starting point and the transfer run (i.e., beginning and end of neurofeedback training, in which no feedback was provided; Fig. 2B). Individuals above the p-Tau181 cut-off significantly downregulated hippocampal activity during neurofeedback but only with feedback from the hippocampus (*p* = .005), not with feedback from the control region (*p* = .647; Supplementary Table 4). In addition, individuals below that cut-off downregulated hippocampal activity with feedback from the hippocampus (*p* = .056) but not with feedback from the control region (*p* = .229). This was similar in individuals with high or low amyloid risk, as they significantly downregulated hippocampal activity when receiving feedback from the hippocampus (high: *p* = .011, low: *p* = .037) but not when receiving feedback from the control region (high: *p* = .249, low: *p* = .409; Supplementary Table 5). Classifying participants according to clinical assessment, feedback from the hippocampus reduced activity in the hippocampus (patients with MCI, *p* = .035; healthy volunteers, *p* = .010), while feedback from the control region did not significantly reduce activity in that region (*p* = .331) or in the hippocampus (*p* = .162; Supplementary Table 6).

### Hippocampal neurofeedback was associated with reduced hippocampal activity one week later

We next tested whether the effects of neurofeedback on hippocampal activity outlasted the neurofeedback session. One week after the last neurofeedback training, all participants performed the fMRI pattern separation task again and hence, task performance as well as hippocampal activity were assessed again. Classifying participants based on p-Tau181 (Fig. 4A; Supplementary Table 7), individuals above or below the cut-off still had significantly reduced hippocampal activity a week after having downregulated hippocampal activity (above: *p* = .009, below: *p* = .011) but not after having downregulated activity in a control region (above: *p* = .553, below: *p* = .565). Classifying participants based on amyloid positivity risk (Fig. 4B; Supplementary Table 8), those with high amyloid risk still had significantly reduced activity in the hippocampus after downregulation of hippocampal activity one week earlier (*p* = .006) but not after downregulation of activity in the control region (*p* = .253). Those will low amyloid risk also had slightly reduced hippocampal activity when downregulating hippocampal activity one week earlier (*p* = .065), but not when regulating the control region (*p* = .908). In addition, both patients with MCI (*p* = .030) and healthy volunteers (*p* = .011) still had significantly reduced activity in the hippocampus, but only if they had downregulated hippocampal activity during neurofeedback. Regulating activity in a control region had no significant effect on hippocampal activity (*p* = .718; Figure 4C, Supplementary Table 9). These results indicate that successful reduction of hippocampal activity depends on feedback from the hippocampus. Successful downregulation worked particularly well in people with high risk of amyloid pathology that predisposes to higher hippocampal activity, even if they had not shown high activity in the hippocampus at baseline.

**Figure 4.**
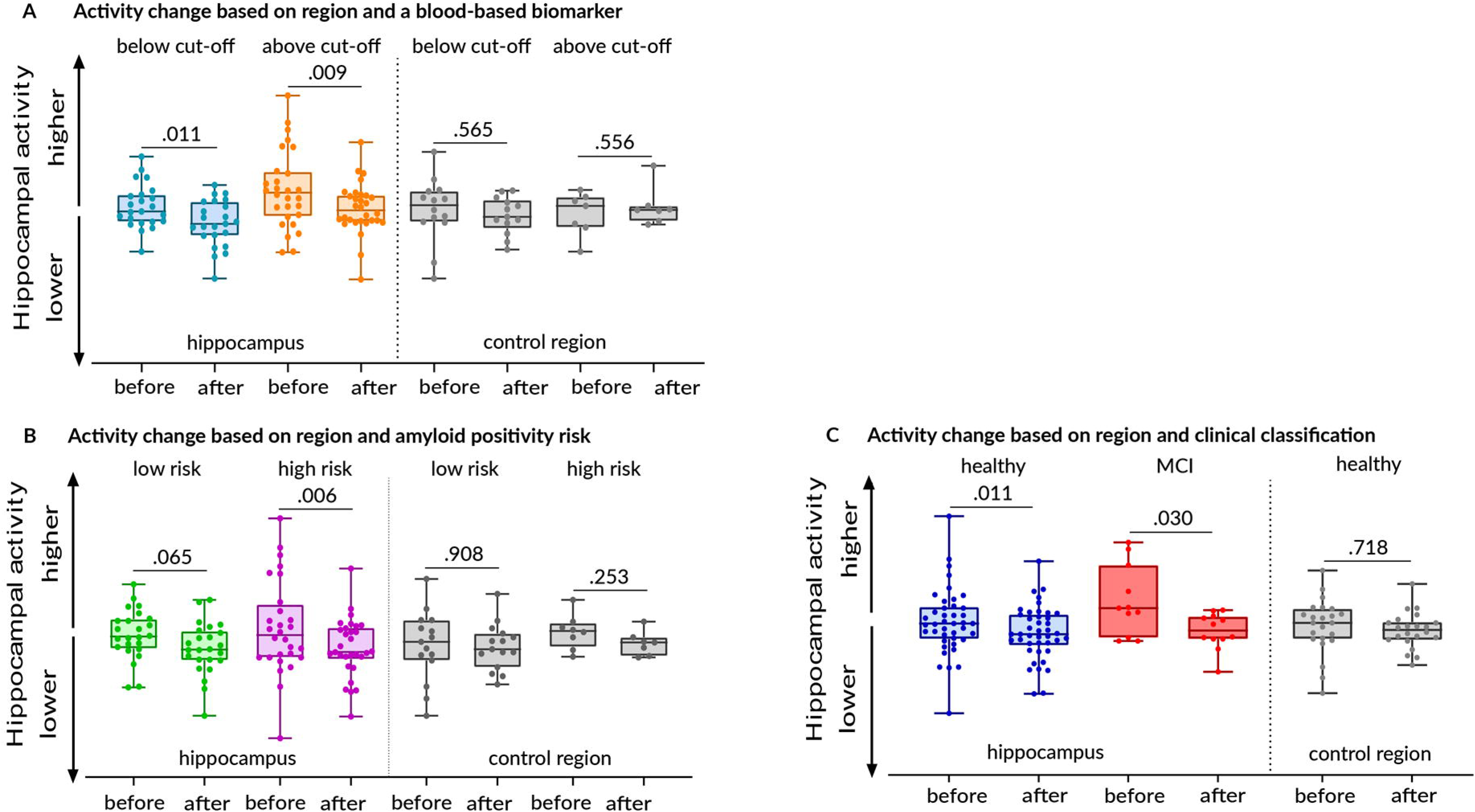
Change in hippocampal activity one week after neurofeedback. We split the sample based on **(A)** a blood-based biomarker, **(B)** amyloid positivity risk calculation, or **(C)** clinical diagnosis. Error bars indicate standard error of the mean. Boxplots indicate min to max values.

### Hippocampal neurofeedback improved pattern separation performance

We next examined whether successful regulation of the hippocampus influenced pattern separation performance, a task closely associated with the hippocampus. To evaluate task performance, we used the lure discrimination index (LDI) that tracks the ability to remember the rich details of items to correctly identify them as ‘similar’ and not endorse them as ‘old’ (23). In addition, we used the number of correctly identified similar items as well as errors (i.e., misidentifications). In individuals above the p-Tau181 cut-off, task performance improved one week after downregulating activity in the hippocampus as they were better able to identify similar items (*p* < .001; Fig. 5A) and made fewer errors (*p* = .004; Supplementary Table 7). This was not the case if they had downregulated activity in the control region, as they did not identify more similar items correctly (*p* = .226) or did not make fewer errors (*p* = .243). For individuals below cut-off, downregulation of activity in the hippocampal or in a control region improved the identification of similar items (hippocampus, *p* = .010; control region, *p* = .013). However, fewer errors were only made if they had downregulated hippocampal activity (*p* = .045) but not if they had downregulated activity in the control region (*p* = .587; Supplementary Table 7). This was similar for individuals with high amyloid risk according to the Palmquist formula, as their task performance improved (*p* = < .001) and they made fewer errors (*p* < .001; Fig. 5B) when they had downregulated activity in the hippocampus but not when downregulating activity in the control region (*p* = .227, errors: *p* = .542; Supplementary Table 8). Individuals with low risk improved their task performance regardless of the brain region they regulated (hippocampus: *p* = .016; control region: *p* = .001). When classifying individuals clinically, all participants improved their task performance if they downregulated activity in the hippocampus (patients with MCI: *p* = .026; healthy participants: *p* < .001) or in a control region (*p* = .001; Fig. 5C). However, errors were only reduced with downregulation of hippocampal activity (patients with MCI: *p* = .020; healthy participants: *p* = .008) but not with downregulation of the control region (*p* = .166; Supplementary Table 9).

**Figure 5.**
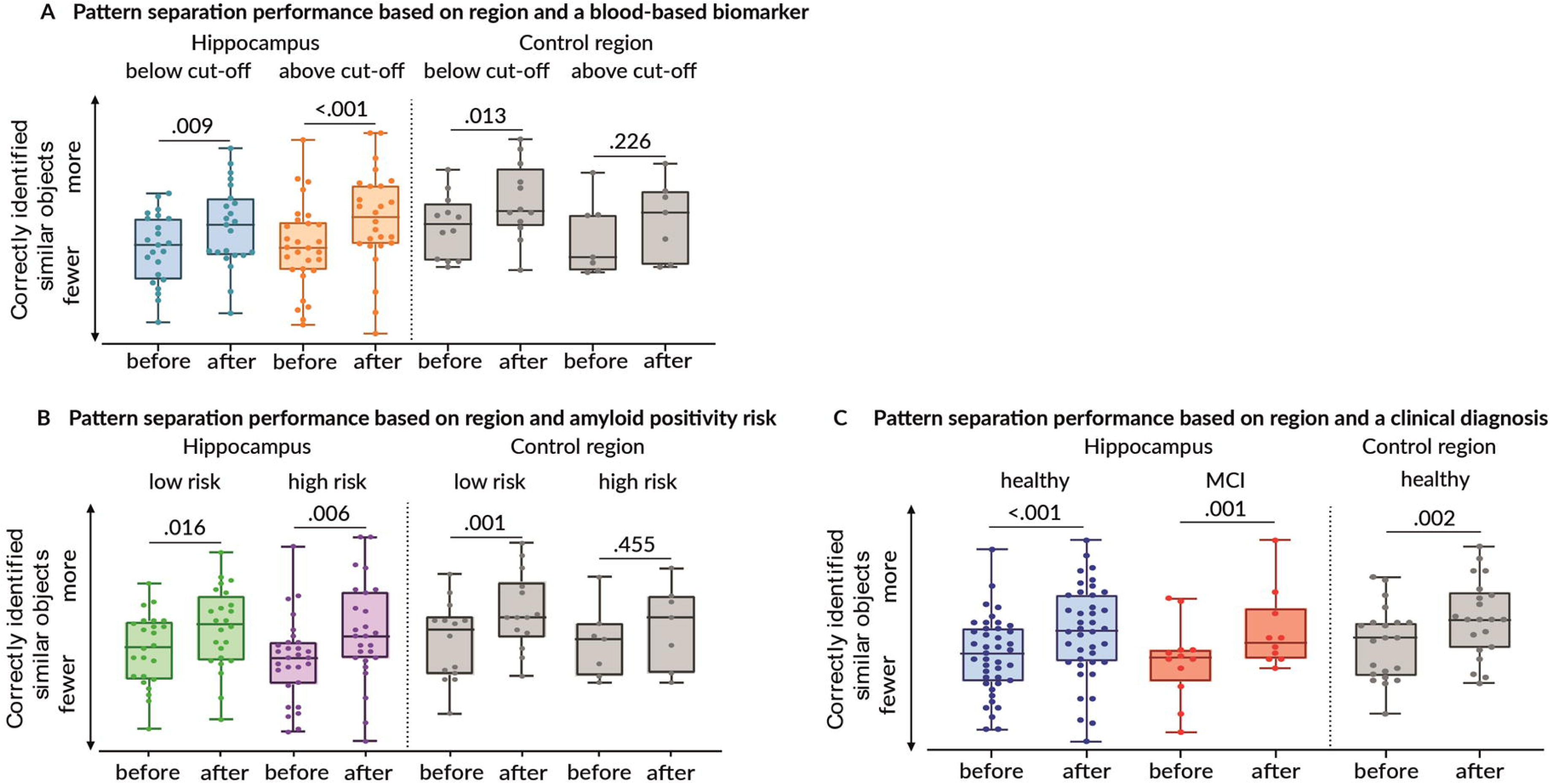
Change in pattern separation performance one week after neurofeedback. We split the sample based on (A) a blood-based biomarker, (B) amyloid positivity risk, or (C) clinical diagnosis. Error bars indicate standard error of the mean. Boxplots indicate min to max values.

These results indicate that individuals with a high risk of developing AD only improved their task performance when they downregulated hippocampal activity, the brain region necessary for this task. This was regardless of whether they initially showed hippocampal hyperactivity before neurofeedback. In individuals with low risk of amyloid pathology, both downregulation of the hippocampus or a control region improved the identification of similar items. However, errors were only reduced when the hippocampus was downregulated during neurofeedback.

### The reduction of hippocampal activity was significantly correlated with pattern separation improvement

We went on to examine whether the amount of hippocampal downregulation related to the amount of improvement in the pattern separation task. In participants above the p-Tau181 cut-off, we found a significant negative correlation after downregulation of hippocampal activity. Stronger downregulation was associated with stronger improvement in the pattern separation task (*p* = .014; Fig. 6A). This was not the case for the control region (*p* = .343). We found a similar result for participants with high amyloid risk after downregulation of hippocampal activity (*p* = .041; Fig. 6B) but not for the control region (*p* = .523). In patients with MCI, stronger downregulation of hippocampal activity was again significantly negatively correlated with better task performance (*p* = .046; Fig. 6C). We found no significant correlation in those below the p-Tau181 cut-off (hippocampus: *p* = .765; control region: *p* = .639) or with low amyloid risk (hippocampus: *p* = .801; control region: *p* = .574), regardless of the region they regulated during neurofeedback. We also found no significant correlation in cognitively healthy volunteers, regardless of the region they regulated (hippocampus, *p =* .429; control region, *p* = .620).

**Figure 6.**
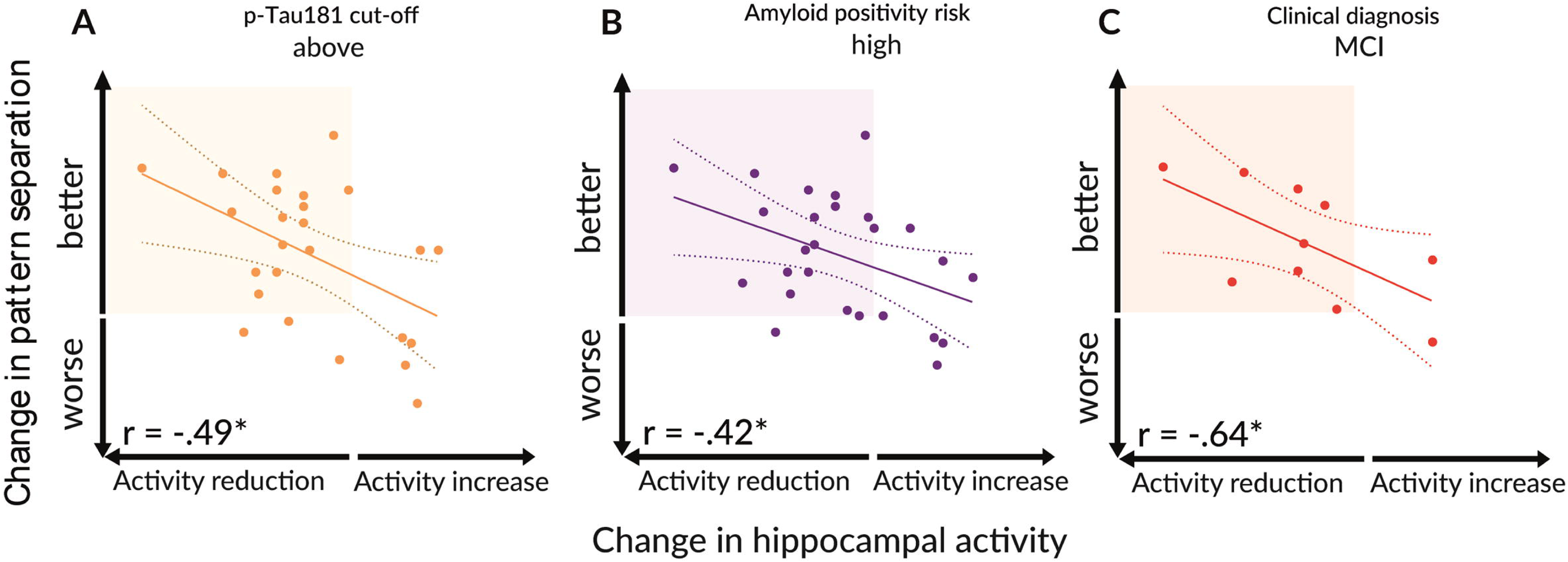
Correlation between change in hippocampal activity and change in pattern separation performance. Participants downregulated activity in the hippocampus and were **(A)** above cut-off in a blood-based biomarker, **(B)** at high risk of amyloid positivity, or **(C)** had a clinical diagnosis of mild cognitive impairment (MCI). Change in activity and performance was calculated using values from before to after neurofeedback. There were approximately 4 weeks between both assessments. Highlighted areas indicate participants that downregulated hippocampal activity and improved their task performance. Significant at *p* < .05*.

## Discussion

The accumulation of amyloid pathology in the brain results in hippocampal hyperactivity, faster disease progression and more pronounced cognitive decline [36]. Hippocampal hyperactivity may thus be detrimental both for cognitive performance and pathological progression. If so, reducing hippocampal hyperactivity promises to improve hippocampal dependent cognition and slow down disease progression. The results of the current study provide evidence to support this assumption. Two sessions of real-time fMRI neurofeedback, one week apart and for 1 hour each, significantly reduced hippocampal activity within the neurofeedback sessions, with the effect still detectable one week later. Importantly, pattern separation performance - a task closely associated with the hippocampus – improved with the effect still detectable one week later. Downregulating activity in a control region had no influence on hippocampal activity or pattern separation performance. Our results were consistent when using three different and independent classification approaches to identify people at an increased risk of amyloid pathology. Individuals above the p-Tau181 cut-off, individuals with high amyloid risk based on a formula, or patients with a clinical diagnosis of amnestic MCI were all able to reduce hippocampal activity and subsequently improved pattern separation task performance – but only when they downregulated hippocampal activity during neurofeedback. The effects were demonstrable a week later but were not seen when participants downregulated a control region. This indicates that performance improvements were specific for regulating activity in the region closely linked to pattern separation, i.e., the hippocampus. Hence, during neurofeedback, participants learned how to solve the pattern separation task with less hippocampal activation. Neurofeedback likely involves operant conditioning. Individuals are rewarded for activity changes with positive feedback (here: shift in colour in thermometer icon from red towards green), which encourages them to reproduce it and helps them learn how to regulate it, which in turn reduces brain activity. Once participants have successfully learned to control their brain activity, a ‘transfer run’ at the end of the training examines whether they can maintain this control without the presence of feedback. Our results indicate that hippocampal activity at the end of the training (i.e., during this ‘transfer run’) was significantly lower than before neurofeedback – but only when the hippocampus was downregulated. The ability to regulate hippocampal activity likely depended on the strategies used during training. The most commonly used strategies were imagination (e.g., visualizing a walk in the forest), active thinking (e.g., mentally reciting the alphabet), focusing on body parts (e.g., concentrating on the feet), or focusing on parts of the stimuli (e.g., a specific part of an image). These strategies indicate that participants, while viewing the images, tried to relax and avoided overly focusing on the stimuli.

The improvement in pattern separation performance with hippocampal downregulation comprised a higher lure discrimination index (LDI), better identification of similar items, and fewer errors. Different sub-regions within the hippocampus contribute to the ability to discriminate similar from old or new stimuli. While the dentate gyrus is responsible for pattern separation, the CA3 region may be important for pattern completion [32]. A balance between pattern separation and pattern completion is crucial for efficient memory retrieval. If shifted too far in one direction, memories that are distinct but related may either be stored as completely separate or as overly similar, impairing the ability to differentiate between them. The dentate gyrus receives input from the perforant pathway, in which synapses are lost during ageing and, even more pronounced, in patients with MCI [37]. This loss of synapses leads to a drastically reduced cortical input to the dentate gyrus and, hence, to an impairment in pattern separation. In the CA3 region, on the other hand, there is reduced inhibition of recurrent collaterals when cholinergic modulation of inhibitory interneurons becomes impaired (Fig. 7A). This leads to hyperactivity, which has frequently been observed in patients with MCI [6]. An over-active CA3 region promotes pattern completion rather than pattern separation and hence, more errors occur in a pattern separation task (Fig. 7B). In our study, only downregulation of hippocampal activity led to a significant reduction of such errors, and particularly so in individuals at risk for amyloid positivity. This indicates that the reduction of hippocampal activity mainly influenced the CA3 region of the hippocampus, most likely by normalising the imbalance between pattern separation and pattern completion (Fig. 7C). Importantly, this did not require hippocampal hyperactivity *before* neurofeedback. We only found significantly increased hippocampal activity (i.e., hyperactivity) when comparing patients with amnestic MCI to healthy individuals but not when comparing individuals at high risk of amyloid positivity to those at low risk. One explanation could be that clinical classification was only possible with more advanced AD pathology. Indeed, medial temporal lobe atrophy and p-Tau181 values were significantly higher in patients with amnestic MCI than in healthy individuals at high (atrophy: *p* = .001; p-Tau: *p* = .022) or low risk of amyloid positivity (atrophy: *p* < .001; p-Tau: *p* < .001) (Fig. 8A and B). There may be a relationship between the amount of amyloid pathology and hippocampal hyperactivity, at least in rodents [38]. Participants at high risk of amyloid positivity based on blood-based biomarkers may have been at an earlier stage in the evolution of AD pathology, and, thus, may not yet be at a stage with overt hippocampal hyperactivity. Still, reducing activity in the hippocampus in those individuals improved their pattern separation performance and, hence, the efficiency of hippocampal function. This indicates that downregulation of hippocampal activity may be beneficial in people at high risk of AD pathology even before evidence of hyperactivity emerges. We show here that neurofeedback, a non-pharmacological intervention, could serve that purpose.

**Figure 7.**
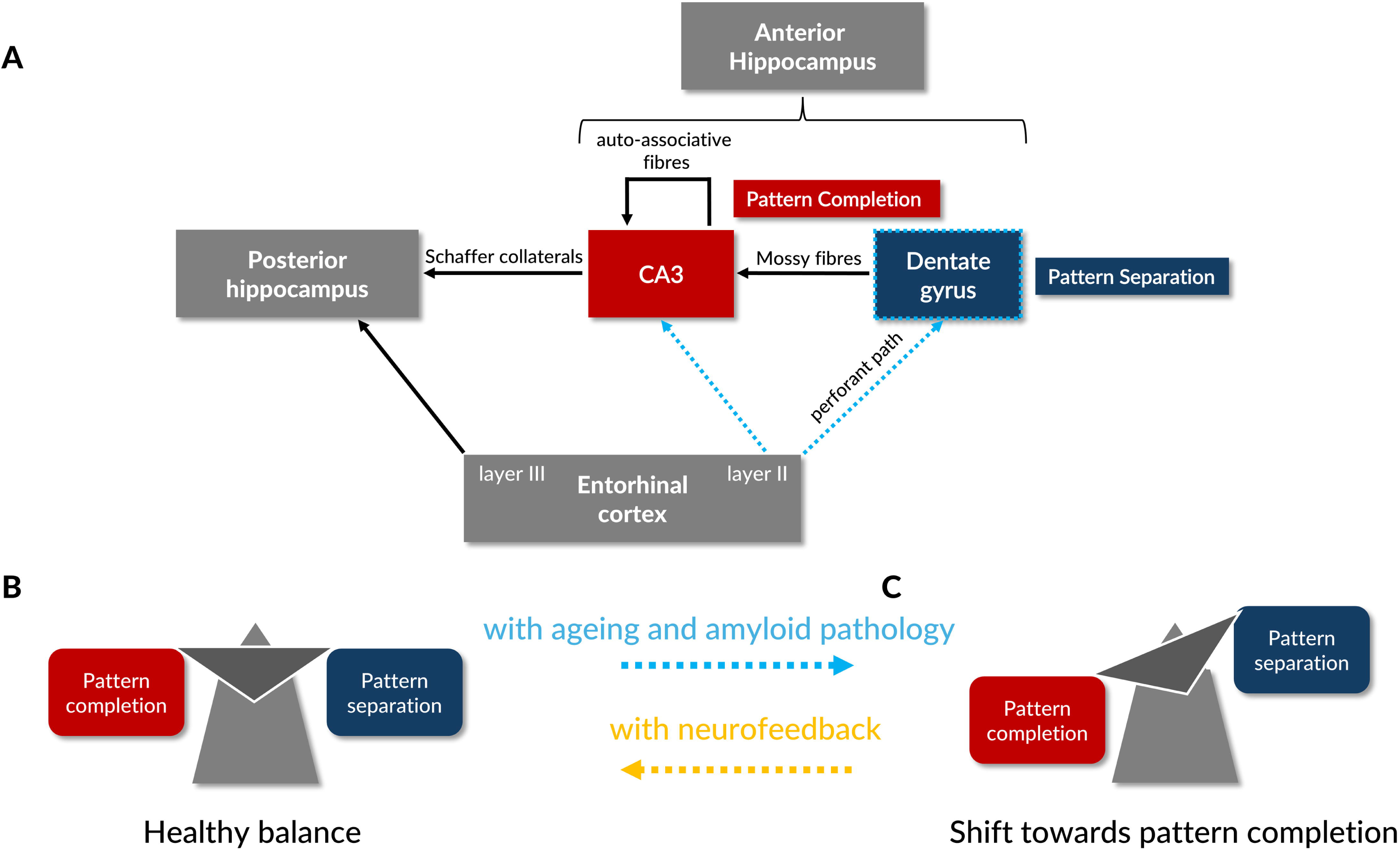
Functional changes in the hippocampus during ageing or with amyloid pathology. **(A)** The entorhinal cortex provides less input to the dentate gyrus and CA3, and the DG is less excitable (depicted with blue dotted lines). These changes may increase CA3 activity through recurrent collaterals (auto-associative fibres) and **(B)** shift the balance between pattern completion and pattern separation towards pattern completion. **(C)** Neurofeedback may reverse that.

**Figure 8.**
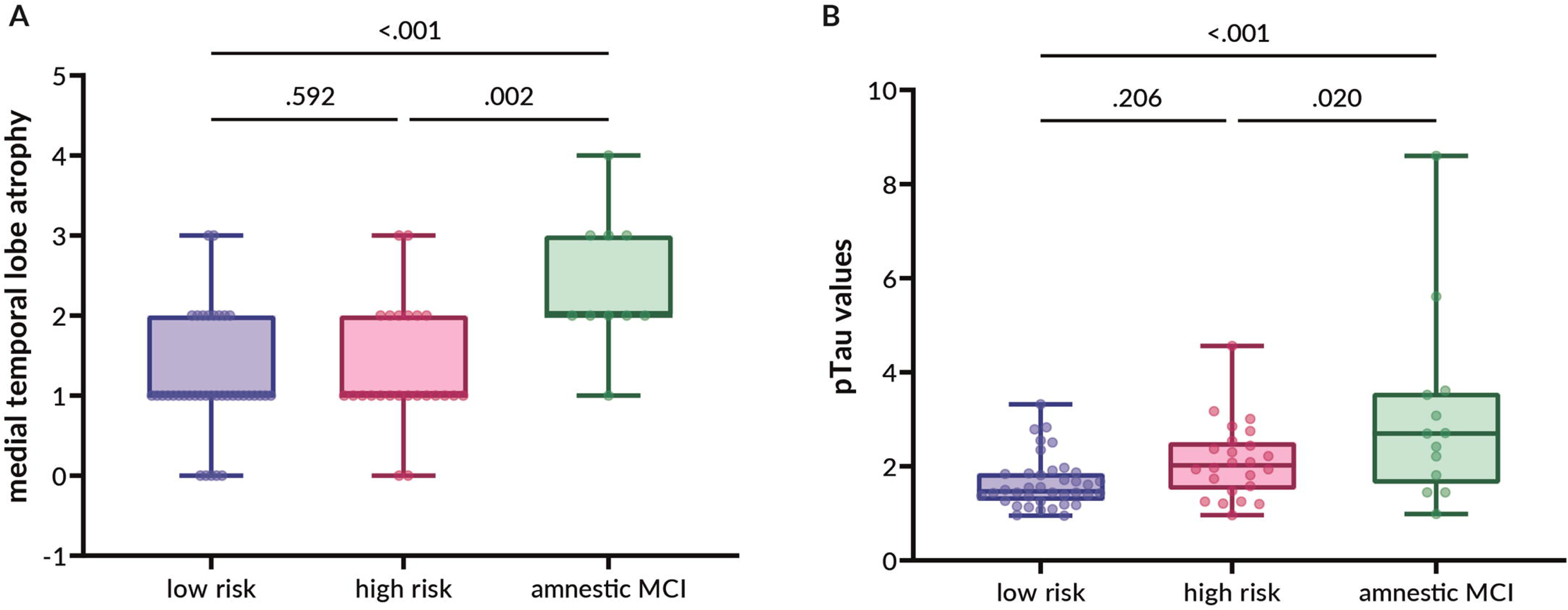
Variables indicating Alzheimer’s disease pathology in individuals at low or high risk of amyloid positivity and in patients with Mild Cognitive Impairment (MCI). We evaluated **(A)** medial temporal lobe atrophy or **(B)** phosphorylated Tau (p-Tau181) in individuals at risk of amyloid positivity. The risk was calculated independently from medial temporal lobe atrophy or p-Tau181 values. Boxplots indicate min to max values.

Other approaches to reduce hippocampal hyperactivity in patients with MCI include levetiracetam, an antiepileptic drug [2,3], which also improved pattern separation performance. Levetiracetam seems to bind specifically to synaptic vesicle protein 2A thereby regulating neurotransmitter release and neuronal (hyper)excitability [39]. While levetiracetam may induce broader, more widespread effects in the brain, fMRI neurofeedback can target region-specific brain activity. In addition, neurofeedback may give participants a sense of empowerment as they themselves regulate activity without having to ‘depend on’ taking medication. However, fMRI neurofeedback requires an MRI scanner. This excludes individuals with contraindications to MRI such as metallic implants or metallic fragments, and in regions where no MRI is available, such an intervention would not be possible. However, the strategies used by the participants to downregulate hippocampal activity may also be valuable without MRI. If these strategies reduced hippocampal activity without neurofeedback, they may be implemented in an app for daily use on a mobile phone.

Taken together, our study shows that reducing hippocampal activity is beneficial in people at increased risk of amyloid positivity even *before* a clinical diagnosis of MCI is made and even *before* there is hippocampal hyperactivity. Hippocampal hyperactivity is thought to contribute to the detrimental effects of progressive Alzheimer pathology on the hippocampus leading to neuronal loss and functional impairment. Our data suggest that a reduction of hippocampal activity is beneficial early in the evolution of AD pathology. If confirmed this argues for a more systematic evaluation of the effects of early downregulation of hippocampal activity including on the evolution of AD pathology. We also show that a non-pharmacological intervention, neurofeedback, can achieve the goal of downregulating hippocampal activity even a week after the neurofeedback session. Neurofeedback may be an additional, complementary, treatment approach. It will be important to examine whether using a drug, such as levetiracetam, in combination with a non-pharmacological treatment such as neurofeedback can produce synergistic effects. This does not address the cause of AD. However, it may contribute to improving hippocampus dependent cognition and slowing down the inevitable progression of AD pathology.

## Limitations

Our study may have some limitations. Real-time fMRI neurofeedback requires a complex methodological setup and an MRI scanner. Hence, individuals with contraindications to MRI (e.g., cardiac pacemakers or cochlear implants) cannot participate and the intervention is restricted to areas where an MRI is available. In a future study, we will examine whether mental strategies that helped participants to downregulate hippocampal activity will lead to similar results. This would allow hippocampal downregulation independent of an MRI scanner.

## Conclusion

We show that reducing hippocampal activity using neurofeedback is beneficial in people at increased risk of amyloid positivity before a clinical diagnosis of MCI is made and even before there is hippocampal hyperactivity. Hence, neurofeedback may be an adaptable intervention that directly targets hippocampal hyperactivity in those at risk of AD.

## Acknowledgements

We thank Andrea Federspiel for his help in the MRI set-up as well as in quality control of pilot data. We thank Arnold Bakker for his input on the study design. We thank Amelie Haugg, Roland Sladky, and Frank Scharnowski for their support in setting up the real-time fMRI neurofeedback pipeline. We thank Saskia Salzmann, Lea Pfister, Loris Rohrbach- Grandjean, Nathalie Schwab, Julia Schmid, Carla Stadelmann, Antonia Rumpf, Claudia Klopfenstein, Mitja Mosimann, Jenny Schnüriger, and Nadine Schmidt for their help in data acquisition. We thank all participants for their time and support of the study.

## Data availability

Raw data were generated at the University of Bern. Derived data supporting the findings of this study are available from the corresponding author on request.

## Consent statement

All participants provided written informed consent.

## Footnotes

1 For the determination of sample size, we used G*power [13]. Since no study so far examined downregulation of hippocampal activity, we opted for the detection of a small effect (i.e., Cohen’s *f* = 0.25). This required an inclusion of n = 84 participants for a repeated measures design with 4 groups and 4 measurements. We used the following criteria to calculate sample size: ANOVA repeated measures within-between interaction, α err = 0.01, 1-β = 0.99, number of groups = 4, number of measurements = 4.

