## Supplementary Material for "Downregulation of hippocampal activity improves memory performance in individuals at risk of Alzheimer’s disease"

### **Supplement**

In this supplement, we first report deviations from our study protocol ^1^ and then we provide full statistics of results reported in the main manuscript.

##### Deviations from our study protocol

We initially planned to include n = 84 participants and to classify them based on clinical characterization (i.e., healthy or patients with MCI). Due to advances in the field of blood based biomarkers ^2,3^ and since hippocampal hyperactivity seems to be related to Alzheimer’s disease (AD) pathology ^4^, we decided to use a different approach and to classify participants based on AD pathology using three classification methods. We also planned to apply a 12-month follow-up assessment. Due to the COVID-19 pandemic, access to MRI facilities was limited. Therefore, data acquisition was delayed and we could not include another MRI assessment for all participants after 12 months. Instead of APOE and BDNF, we used hippocampal volume as a covariate in our statistical model. Hippocampal volume is a more meaningful covariate since it directly relates to hippocampal activity.

**Table S1.** Characterisation of study participants when divided by p-Tau181 cut-off for amyloid positivity before our intervention took place (mean ± standard deviation).

|  |  | Above cut-off |  | Below cut-off | *p*-value |
| --- | --- | --- | --- | --- | --- |
| Number (f/m) |  | 38 (23/15) |  | 37 (22/15) | .925 |
| Age in years |  | 70.21 ± 5.80 |  | 68.32 ± 6.09 | .174 |
| Education in years |  | 16.03 ± 2.43 |  | 14.97 ± 2.08 | .053 |
| Total intracranial volume in cm^3^/ml |  | 1473.54 ± 129.31 |  | 1504.42 ± 149.54 | .916 |
| Hippocampal volume in %TIV |  | 0.23 ± 0.04 |  | 0.26 ± 0.02 | **.001** |
| Hippocampal activity |  | 0.02 ± 0.26 |  | -0.08 ± 0.18 | .086 |
| Medial temporal lobe atrophy (left; 0-4) |  | 1.68 ± 0.95 |  | 1.14 ± 0.69 | **.009** |
| Medial temporal lobe atrophy (right; 0-4) |  | 1.44 ± 0.89 |  | 1.23 ± 0.65 | .260 |
| Lure discrimination index |  | 0.23 ± 0.23 |  | 0.32 ± 0.13 | **.044** |
| Verbal delayed recall |  | 8.58 ± 3.85 |  | 10.95 ± 3.06 | **.004** |

*Abbreviations*: f/m = female/male, cm = centimetre, ml = millilitre, TIV = total intracranial volume.

**Table S2.** Characterisation of study participants when divided by amyloid positivity risk (high or low) before our intervention took place (mean ± standard deviation). Significant differences after Bonferroni correction are shown in bold.

|  |  | High risk |  | Low risk | *p*-value |
| --- | --- | --- | --- | --- | --- |
| Number (f/m) |  | 39 (23/16) |  | 39 (25/14) | .642 |
| Age in years |  | 70.64 ± 6.16 |  | 68.05 ± 5.58 | .062 |
| Education in years |  | 15.44 ± 2.44 |  | 15.42 ± 2.19 | .965 |
| Total intracranial volume in cm^3^/ml |  | 1479.04 ± 148.52 |  | 1485.40 ± 135.59 | .844 |
| Hippocampal volume in %TIV |  | 0.23 ± 0.04 |  | 0.26 ± 0.03 | **.003** |
| Hippocampal activity |  | -0.01 ± 0.28 |  | -0.08 ± 0.19 | .227 |
| Medial temporal lobe atrophy (left; 0-4) |  | 1.65 ± 0.92 |  | 1.18 ± 0.73 | **.020** |
| Medial temporal lobe atrophy (right; 0-4) |  | 1.44 ± 0.89 |  | 1.26 ± 0.69 | .343 |
| Lure discrimination index |  | 0.23 ± 0.18 |  | 0.33 ± 0.14 | **.006** |
| Verbal delayed recall |  | 8.03 ± 3.53 |  | 11.41 ± 2.83 | **< .001** |

*Abbreviations*: f/m = female/male, cm = centimetre, ml = millilitre, TIV = total intracranial volume.

**Table S3.** Characterisation of study participants when divided by clinical diagnosis (i.e., Mild Cognitive Impairment or healthy older volunteers) before our intervention took place (mean ± standard deviation). Significant differences after Bonferroni correction are shown in bold.

|  |  | MCI |  | Healthy volunteers |  |  | *p*-value |
| --- | --- | --- | --- | --- | --- | --- | --- |
| Number (f/m) |  | 13 (4/9) |  | 65 (44/21) |  |  | .012 |
| Age in years |  | 71.66 ± 5.05 |  | 68.85 ± 6.06 |  |  | .136 |
| Education in years |  | 16.83 ± 2.62 |  | 15.16 ± 2.15 |  |  | .020 |
| Total intracranial volume in cm^3^/ml |  | 1521.92 ± 117.16 |  | 1474.29 ± 145.14 |  |  | .270 |
| Hippocampal volume in %TIV |  | 0.21 ± 0.03 |  | 0.26 ± 0.03 |  |  | **< .001** |
| Hippocampal activity |  | 0.10 ± 0.28 |  | -0.07 ± 0.23 |  |  | **.033** |
| Medial temporal lobe atrophy (left; 0-4) |  | 2.40 ± 0.83 |  | 1.24 ± 0.74 |  |  | **< .001** |
| Medial temporal lobe atrophy (right; 0-4) |  | 1.90 ± 1.10 |  | 1.26 ± 0.70 |  |  | **.016** |
| Lure discrimination index |  | 0.10 ± 0.26 |  | 0.32 ± 0.15 |  |  | **< .001** |
| Verbal delayed recall |  | 5.31 ± 2.95 |  | 10.64 ± 3.05 |  |  | **< .001** |

*Abbreviations*: f/m = female/male, cm = centimetre, ml = millilitre, TIV = total intracranial volume.

**Table S4.** Change in brain activity from starting point to transfer run in individuals receiving neurofeedback from the hippocampus or a control region. Participants were divided by p-Tau181 cut-off for amyloid positivity (below or above) and the region they regulated during neurofeedback.

|  |  | Starting point | Transfer run | *F* | df | *p* | *β* | 95% CI |
| --- | --- | --- | --- | --- | --- | --- | --- | --- |
| **Above cut-off - hippocampus** | |  |  |  |  |  |  |  |
| Hippocampal activity | | 0.13 ± 0.13 | -0.003 ± 0.20 | 9.03 | 1,29 | **.005** | -0.13 | -0.22, -0.04 |
| **Below cut-off - hippocampus** | |  |  |  |  |  |  |  |
| Hippocampal activity | | 0.12 ± 0.11 | 0.06 ± 0.12 | 4.07 | 1,22 | .056 | -0.06 | -0.12, 0.005 |
| **Above cut-off – control region** | |  |  |  |  |  |  |  |
| Hippocampal activity  IPS activity | | 0.20 ± 0.18  0.06 ± 0.47 | 0.15 ± 0.33  0.24 ± 0.45 | 0.23  1.16 | 1,6  1,6 | .647  .293 | -0.04  0.18 | -0.24, 0.16  -0.16, 0.52 |
| **Below cut-off – control region** | |  |  |  |  |  |  |  |
| Hippocampal activity  IPS activity | | 0.11 ± 0.13  0.33 ± 0.46 | 0.05 ± 0.11  0.12 ± 0.48 | 1.59  3.12 | 1,13  1,13 | .229  .085 | -0.06  -0.21 | -0.16, 0.04  -0.45, 0.03 |

*Abbreviations:* df = degrees of freedom, CI = confidence interval, IPS = regions surrounding the intraparietal sulcus.

**Table S5.** Change in brain activity from starting point to transfer run in individuals receiving neurofeedback from the hippocampus or a control region. Participants were divided by high or low amyloid risk and the region they regulated during neurofeedback. Significant differences after Bonferroni correction are shown in bold.

|  |  | Starting point | Transfer run | *F* | df | *p* | *β* | 95% CI |
| --- | --- | --- | --- | --- | --- | --- | --- | --- |
| **High amyloid risk - hippocampus** | |  |  |  |  |  |  |  |
| Hippocampal activity | | 0.13 ± 0.13 | 0.03 ± 0.18 | 7.32 | 1,30 | **.011** | -0.11 | -0.19, -0.03 |
| **Low amyloid risk - hippocampus** | |  |  |  |  |  |  |  |
| Hippocampal activity | | 0.08 ± 0.13 | 0.004 ± 0.15 | 4.91 | 1,23 | **.037** | -0.08 | -0.16, -0.007 |
| **High amyloid risk – control region** | |  |  |  |  |  |  |  |
| Hippocampal activity  IPS activity | | 0.13 ± 0.03  0.01 ± 0.46 | 0.02 ± 0.20  0.09 ± 0.33 | 1.63  0.23 | 1,6  1,6 | .249  .638 | -0.10  0.07 | -0.28, 0.08  -0.25, 0.39 |
| **Low amyloid risk – control region** | |  |  |  |  |  |  |  |
| Hippocampal activity  IPS activity | | 0.16 ± 0.20  0.38 ± 0.44 | 0.12 ± 0.21  0.21 ± 0.51 | 0.72  2.18 | 1,14  1,14 | .409  .147 | -0.04  -0.17 | -0.14, 0.06  -0.40, 0.06 |

*Abbreviations:* df = degrees of freedom, CI = confidence interval, IPS = regions surrounding the intraparietal sulcus.

**Table S6.** Change in brain activity from starting point and transfer run in individuals receiving neurofeedback from the hippocampus or a control region. Participants were divided by clinical diagnosis (i.e., Mild Cognitive Impairment or healthy volunteers) and the region they regulated during neurofeedback. Significant differences after Bonferroni correction are shown in bold.

|  | Starting point | Transfer run | *F* | df | *p* | *β* | 95% CI |
| --- | --- | --- | --- | --- | --- | --- | --- |
| **Patient with MCI**  Hippocampal activity | 0.16 ± 0.14 | 0.01 ± 0.17 | 5.81 | 1,10 | **.035** | -0.15 | -0.28, -0.02 |
| **Healthy volunteers - hippocampus**  Hippocampal activity | 0.11 ± 0.12 | 0.02 ± 0.17 | 7.38 | 1,41 | **.010** | -0.08 | -0.15, -0.02 |
| **Healthy volunteers – control region**  Hippocampal activity  IPS activity | 0.15 ± 0.16  0.26 ± 0.47 | 0.09 ± 0.20  0.17 ± 0.46 | 2.10  0.96 | 1,20  1,20 | .162  .331 | -0.06  -0.09 | -0.15, 0.02  -0.28, 0.10 |

*Abbreviations:* MCI = Mild cognitive impairment, df = degrees of freedom, CI = confidence interval, IPS = regions surrounding the intraparietal sulcus.

**Table S7.** Brain activity and task performance before and after 2 sessions of neurofeedback in individuals receiving neurofeedback from the hippocampus or a control region. Participants were divided by p-Tau181 cut-off (above or below) and the region they regulated during neurofeedback.

|  | Before | After | *F* | df | *p* | *β* | 95% CI |
| --- | --- | --- | --- | --- | --- | --- | --- |
| **Above cut-off – hippocampus**  Hippocampal activity  Lure discrimination index  Similar items correct (in %)  Similar items incorrect (in %) | 0.05 ± 0.28  0.23 ± 0.20  38.10 ± 17.05  43.26 ± 18.28 | -0.08 ± 0.18  0.37 ± 0.22  51.94 ± 19.21  32.79 ± 17.29 | 7.83  14.10  22.02  10.07 | 1,28  1,28  1,28  1,28 | **.009**  **.001**  **< .001**  **.004** | -0.13  0.13  13.84  -10.47 | -0.22, -0.04  0.06, 0.20  7.92, 19.75  -17.09, -3.85 |
| **Below cut-off – hippocampus**  Hippocampal activity  Lure discrimination index  Similar items correct (in %)  Similar items incorrect (in %) | -0.07 ± 0.16  0.29 ± 0.13  38.94 ± 14.36  46.32 ± 13.48 | -0.19 ± 0.17  0.41 ± 0.17  47.98 ± 16.89  39.99 ± 13.63 | 7.09  7.92  8.13  5.51 | 1,22  1,22  1,22  1,22 | **.011**  **.010**  **.009**  **.045** | -0.12  0.11  9.03  -6.33 | -0.22, -0.03  0.03, 0.19  2.64, 15.43  -12.35, -0.31 |
| **Above cut-off – control region**  Hippocampal activity  Lure discrimination index  Similar items correct (in %)  Similar items incorrect (in %) | -0.12 ± 0.16  0.33 ± 0.21  44.63 ± 15.04  43.32 ± 15.42 | -0.07 ± 0.14  0.45 ± 0.17  50.20 ± 17.79  50.95 ± 15.99 | 0.39  3.58  1.82  0.43 | 1,6  1,6  1,6  1,6 | .556  .107  .226  .542 | 0.05  0.12  7.64  -5.02 | -0.14, 0.24  -0.02, 0.26  -4.97, 20.25  -22.71, 12.67 |
| **Below cut-off – control region**  Hippocampal activity  Lure discrimination index  Similar items correct (in %)  Similar items incorrect (in %) | -0.09 ± 0.23  0.39 ± 0.11  48.10 ± 12.94  36.69 ± 11.58 | -0.13 ± 0.13  0.53 ± 0.15  58.32 ± 15.71  34.23 ± 12.81 | 0.35  12.58  8.81  0.31 | 1,11  1,11  1,11  1,11 | .565  .**005**  **.013**  .587 | -0.04  0.14  10.22  -2.47 | -0.17, 0.09  0.06, 0.23  3.04, 17.41  -11.68, 6.74 |

*Abbreviations:* df = degrees of freedom, CI = confidence interval.

**Table S8.** Brain activity and task performance before and after 2 sessions of neurofeedback in individuals receiving neurofeedback from the hippocampus or a control region. Participants were divided by high or low amyloid risk and the region they regulated during neurofeedback. Significant differences after Bonferroni correction are shown in bold.

|  | Before | After | *F* | df | *p* | *β* | 95% CI |
| --- | --- | --- | --- | --- | --- | --- | --- |
| **High risk – hippocampus**  Hippocampal activity  Lure discrimination index  Similar items correct (in %)  Similar items incorrect (in %) | 0.01 ± 0.31  0.21 ± 0.18  36.34 ± 15.95  46.43 ± 16.95 | -0.13 ± 0.19  0.34 ± 0.22  49.65 ± 19.31  34.84 ± 17.29 | 8.87  11.19  26.94  16.18 | 1,27  1,27  1,27  1,27 | **.006**  **.002**  **< .001**  **< .001** | -0.13  0.12  13.31  -11.59 | -0.22, -0.04  0.05, 0.19  8.17, 18.45  -17.35, -5.82 |
| **Low risk – hippocampus**  Hippocampal activity  Lure discrimination index  Similar items correct (in %)  Similar items incorrect (in %) | -0.06 ± 0.19  0.33 ± 0.15  41.44 ± 14.74  41.40 ± 14.94 | -0.15 ± 0.21  0.45 ± 0.14  50.50 ± 16.20  37.18 ± 14.19 | 3.76  11.35  6.82  1.69 | 1,23  1,23  1,23  1,23 | .065  .**003**  **.016**  .206 | -0.09  0.12  9.06  -4.22 | -0.19, 0.003  0.05, 0.19  2.07, 16.05  -10.76, 2.32 |
| **High risk – control region**  Hippocampal activity  Lure discrimination index  Similar items correct (in %)  Similar items incorrect (in %) | -0.06 ± 0.08  0.36 ± 0.18  44.63 ± 15.04  42.95 ± 12.93 | -0.10 ± 0.09  0.44 ± 0.19  50.20 ± 17.79  37.93 ± 15.03 | 1.65  1.90  0.66  0.43 | 1,5  1,5  1,5  1,5 | .253  .227  .455  .542 | -0.03  0.08  5.58  -5.02 | -0.08, 0.02  -0.06, 0.22  -10.31, 21.47  -22.71, 12.67 |
| **Low risk – control region**  Hippocampal activity  Lure discrimination index  Similar items correct (in %)  Similar items incorrect (in %) | -0.12 ± 0.24  0.35 ± 0.15  44.85 ± 15.59  39.97 ± 13.41 | -0.13 ± 0.16  0.51 ± 0.15  56.67 ± 15.26  35.26 ± 11.97 | 0.01  18.68  16.64  1.65 | 1,13  1,13  1,13  1,13 | .908  **.001**  **.001**  .222 | -0.003  0.16  11.82  -4.72 | -0.15, 0.14  0.08, 0.24  5.84, 17.80  -12.30, 2.87 |

*Abbreviations:* df = degrees of freedom, CI = confidence interval.

**Table S9.** Brain activity and task performance before and after 2 sessions of neurofeedback in individuals receiving neurofeedback from the hippocampus or a control region. Participants were divided by clinical diagnosis (i.e., Mild Cognitive Impairment or healthy volunteers) and the region they regulated during neurofeedback. Significant differences after Bonferroni correction are shown in bold.

|  | Before | After | *F* | df | *p* | *β* | 95% CI |
| --- | --- | --- | --- | --- | --- | --- | --- |
| **Patients with MCI – hippocampus**  Hippocampal activity  Lure discrimination index  Similar items correct (in %)  Similar items incorrect (in %) | 0.10 ± 0.26  0.15 ± 0.20  34.46 ± 14.24  47.90 ± 17.65 | -0.13 ± 0.13  0.30 ± 0.15  52.13 ± 15.66  32.90 ±16.73 | 6.59  6.63  18.13  7.44 | 1,10  1,11  1,11  1,11 | **.030**  **.026**  **.001**  **.020** | -0.23  0.15  17.67  -15.00 | -0.41, -0.04  0.03, 0.27  9.02, 26.33  -26.48, -3.53 |
| **Healthy volunteers – hippocampus**  Hippocampal activity  Lure discrimination index  Similar items correct (in %)  Similar items incorrect (in %) | -0.05 ± 0.25  0.30 ± 0.15  39.79 ± 15.80  43.13 ± 15.74 | -0.14 ± 0.20  0.41 ±0.21  49.43 ± 18.54  36.73 ± 15.74 | 7.11  15.67  16.43  7.79 | 1,41  1,41  1,41  1,41 | **.011**  **< .001**  **< .001**  **.008** | -0.09  0.11  9.63  -6.40 | -0.16, -0.02  0.06, 0.17  4.90, 14.36  -10.97, -1.84 |
| **Healthy volunteers – control region**  Hippocampal activity  Lure discrimination index  Similar items correct (in %)  Similar items incorrect (in %) | -0.10 ± 0.20  0.35 ± 0.16  44.78 ± 15.03  40.86 ± 13.00 | -0.12 ± 0.14  0.49 ± 0.16  54.73 ± 15.87  36.06 ± 12.61 | 0.13  18.61  12.07  2.08 | 1,19  1,19  1,19  1,19 | .718  **< .001**  **.002**  .166 | -0.02  0.14  9.95  -4.81 | -0.12, 0.08  0.07, 0.20  4.14, 15.75  -11.57, 1.96 |

*Abbreviations:* MCI = Mild cognitive impairment, df = degrees of freedom, CI = confidence interval

**Figure S1. Inverted U-shaped model of hippocampal activity as well as trajectory of hippocampal volume in the progression to Alzheimer’s disease.** Figure adapted from Corriveau-Lecavalier et al. ^5^. In a preclinical phase, beginning neurodegenerative processes lead to neuronal hyperactivity. In this time window (highlighted in yellow), interventions to decrease hyperactivity may be important to suspend further activity increases. In later clinical stages, advanced neurodegeneration leads to neuronal hypoactivity.

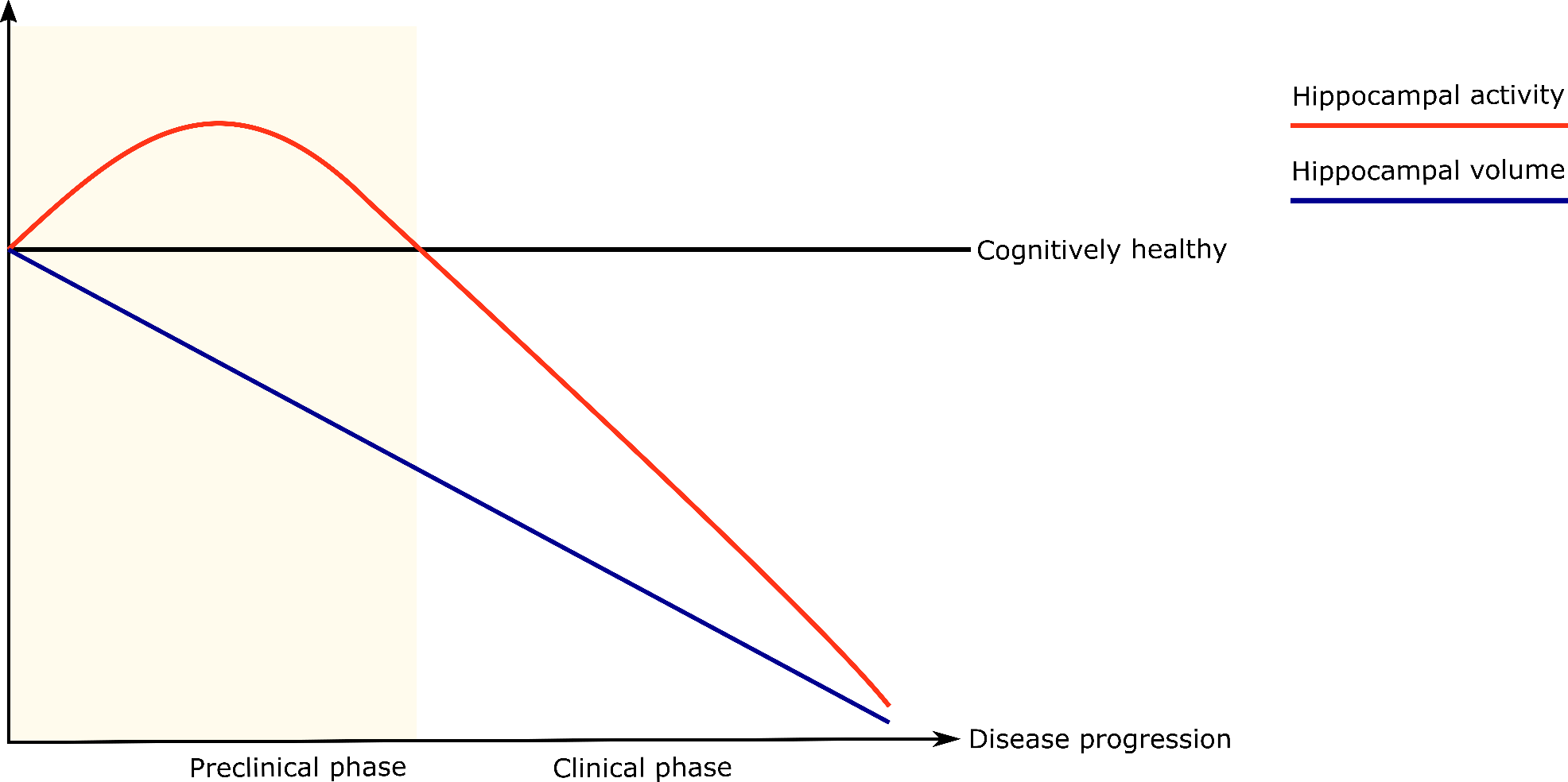

**Figure S2.** Study flow of participants included in our study split by a blood-based biomarker (i.e., p-Tau181). Note that blood was only available after study inclusion.
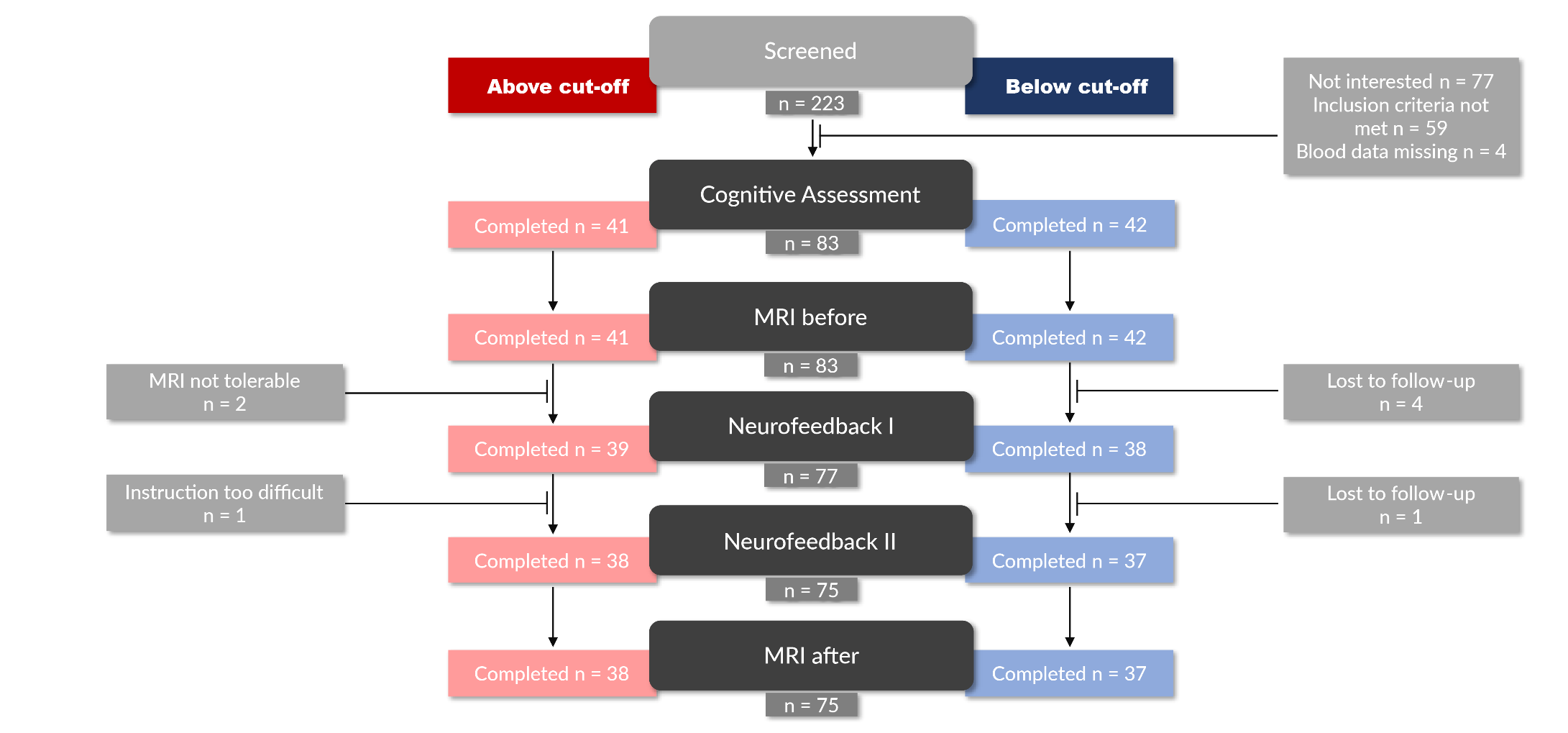

**Figure S3.** Study flow of participants included in our study when split by amyloid positivity risk calculation. Note that a risk calculation was only possible after study inclusion.

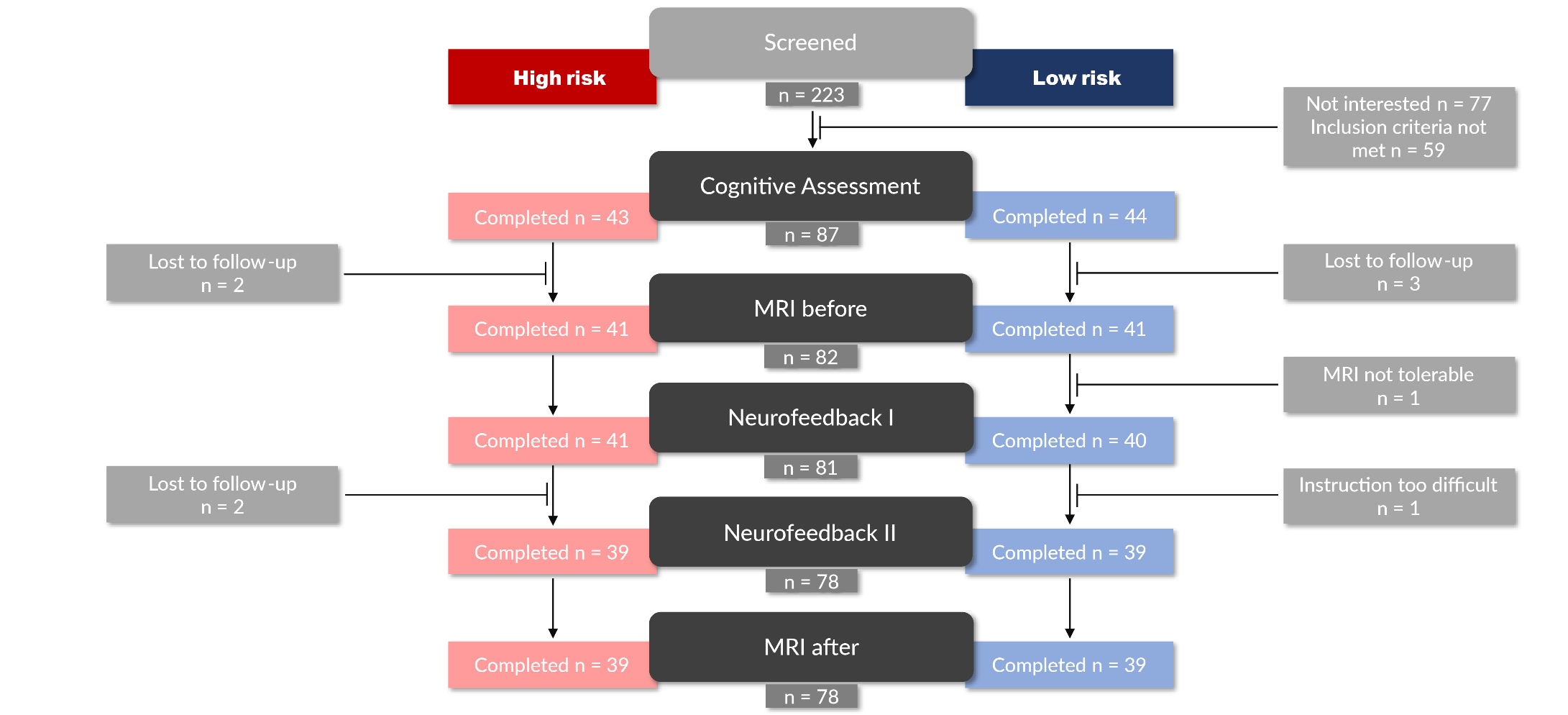

**Figure S4.** Study flow of participants included in our study when split by clinical diagnosis.

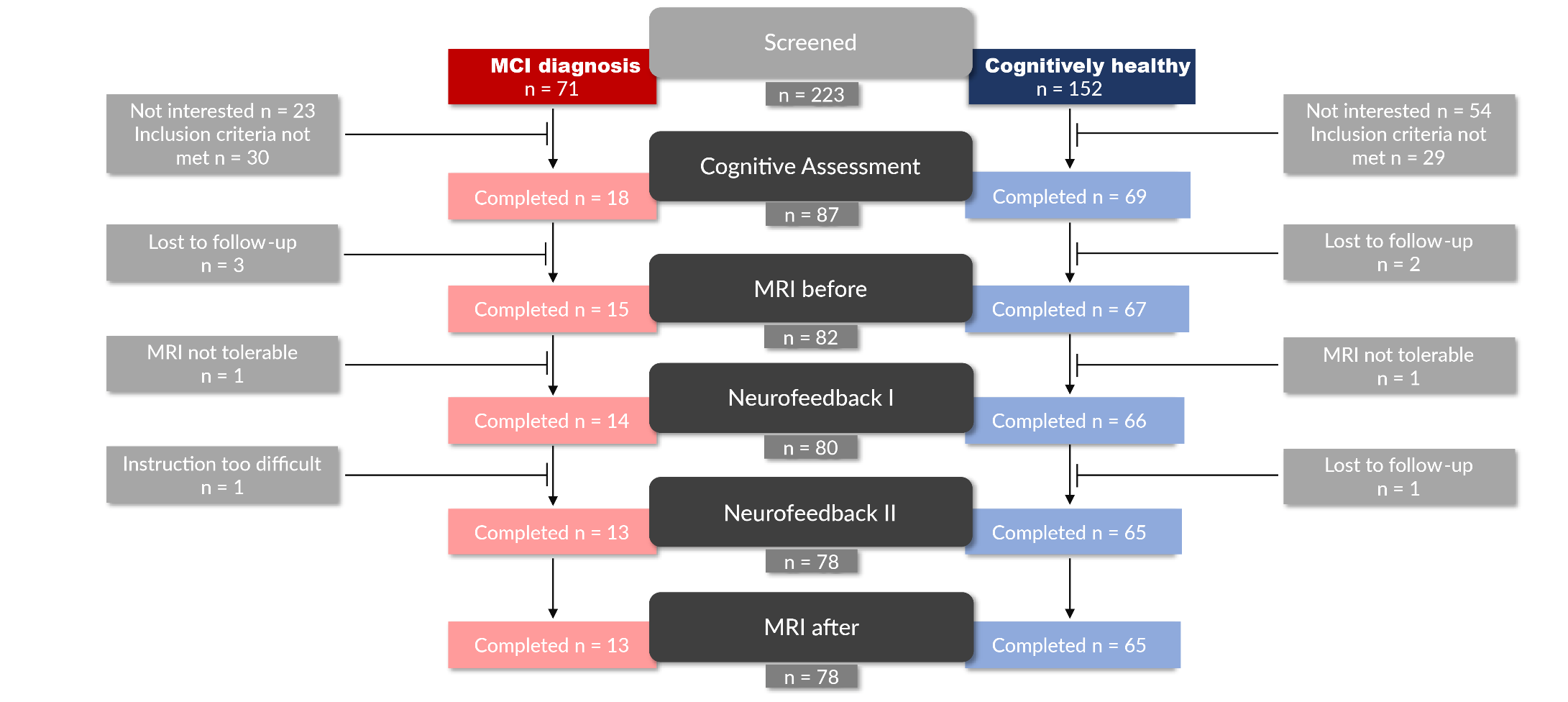
